# Magnitude of Potential Biases in COVID-19 Vaccine Effectiveness Studies due to Differential Healthcare seeking following Home Testing: Implications for Test Negative Design Studies

**DOI:** 10.1101/2024.12.30.24319700

**Authors:** Saba A. Qasmieh, Jill M. Ferdinands, Jessie R. Chung, Ryan E. Wiegand, Brendan Flannery, Madhura S. Rane, Denis Nash

## Abstract

The test-negative design (TND) is widely used to estimate COVID-19 vaccine effectiveness (VE). Biased estimates of VE may result from effects of at-home SARS-CoV-2 rapid diagnostic test (RDT) results on decisions to seek healthcare. To investigate magnitude of potential bias, we constructed decision trees with input probabilities obtained from longitudinal surveys of U.S. adults between March 2022 – October 2023. Prevalence of at-home RDT use and healthcare seeking following a positive or negative RDT result was estimated by participant vaccination status and socio-demographic characteristics. At true *VE* values ranging from 5% to 95%, we defined bias as the difference between the observed and true *VE*. Among 1,918 symptomatic adults, prevalence of at-home RDT use was higher among vaccinated (37%) versus unvaccinated (22%) participants. At-home RDT use was associated with seeking care, and participants reporting positive RDT were more likely than those reporting negative RDT to have sought care when ill. In primary analyses, we observed downward bias in VE estimates that increased in magnitude when true *VE* was low. Variations in proportions of vaccination, at-home RDT use and healthcare seeking by socio-demographic characteristics may impact VE estimates. Further evaluation of potential impact of at-home RDT use on VE estimates is warranted.

## Introduction

The effectiveness of COVID-19 vaccines in protecting against severe outcomes has been largely informed by retrospective observational studies, such as test-negative design (TND) case-control studies^1–3^. Studies that use the TND approach for estimating vaccine effectiveness (VE) aim to reduce bias associated with differential healthcare seeking between vaccinated and unvaccinated persons^4,5^. In such studies, COVID-19 vaccination status is compared between symptomatic patients who test positive for SARS-CoV-2 infection at healthcare facilities, and patients with similar symptoms who test negative for SARS-CoV-2. The design assumes that healthcare seeking is independent of infection status among vaccinated and unvaccinated individuals. When case status is assigned based on testing performed for clinical decision making, uncontrolled factors that influence healthcare seeking may introduce bias if clinical testing varies by patient vaccination and infection status^5–7^. With widespread availability of rapid diagnostic tests for SARS-CoV-2 infection, the effect of self-testing prior to seeking care has the potential to introduce selection bias in TND studies.

The development and approval of rapid SARS-CoV-2 diagnostic tests for home use served a critical public health function for early diagnosis of COVID-19 to mitigate spread of infection and helped alleviate burden on SARS-CoV-2 testing facilities during pandemic waves or increased COVID-19 activity^8,9^. To assess the potential for bias in TND studies of COVID-19 VE among individuals seeking care for COVID-19-like illness, we constructed a decision tree model that simulates a TND case-control study of VE. Informed by a directed acyclic graph (DAG), we explored potential bias in observed VE estimates. We used a simulation model and observational survey data on prevalence of COVID-19 vaccination, use of at-home SARS-CoV-2 rapid diagnostic tests (RDT) and healthcare seeking for COVID-19-like illness among adults ≥ 18 years participating in a diverse U.S. cohort study between March 2022 and October 2023 when rapid at-home COVID-19 test kits were widely available.

## Methods

### Theoretical Basis

Figure 1 presents a DAG depicting potential causal relationships in a TND study of COVID-19 VE. In the assumed causal relationship between COVID-19 vaccination (V_cov_) and COVID-19-like illness (*I_cov_)*, the graph illustrates other potential determinants of V_cov_ and *I_cov_* such as use of at-home *RDT,* home test result (positive or negative), and healthcare seeking behavior (*HS)*. A TND study aims to reduce bias associated with healthcare seeking behavior *HS* by restricting the study population to patients who seek care and receive a diagnostic SARS-CoV-2 test (*T_cov_*). The relationship between *RDT,* V_cov_, and *HS* creates a back-door path between *V_cov_* and *I_cov_*. Selection bias would result if *RDT* and receiving a diagnostic SARS-CoV-2 test with a provider differ by COVID-19 vaccination status, such that vaccinated and unvaccinated patients have different probabilities of selection *T_cov_*(S) into TND following their at-home RDT result. Since the uptake of SARS-CoV-2 home testing has been observed to increase during surges^10^ with differential use among individuals across socio-demographic characteristics^10^, simulations based on survey data may be used to assess the magnitude of potential bias in COVID-19 VE estimates.

**Figure 1:**
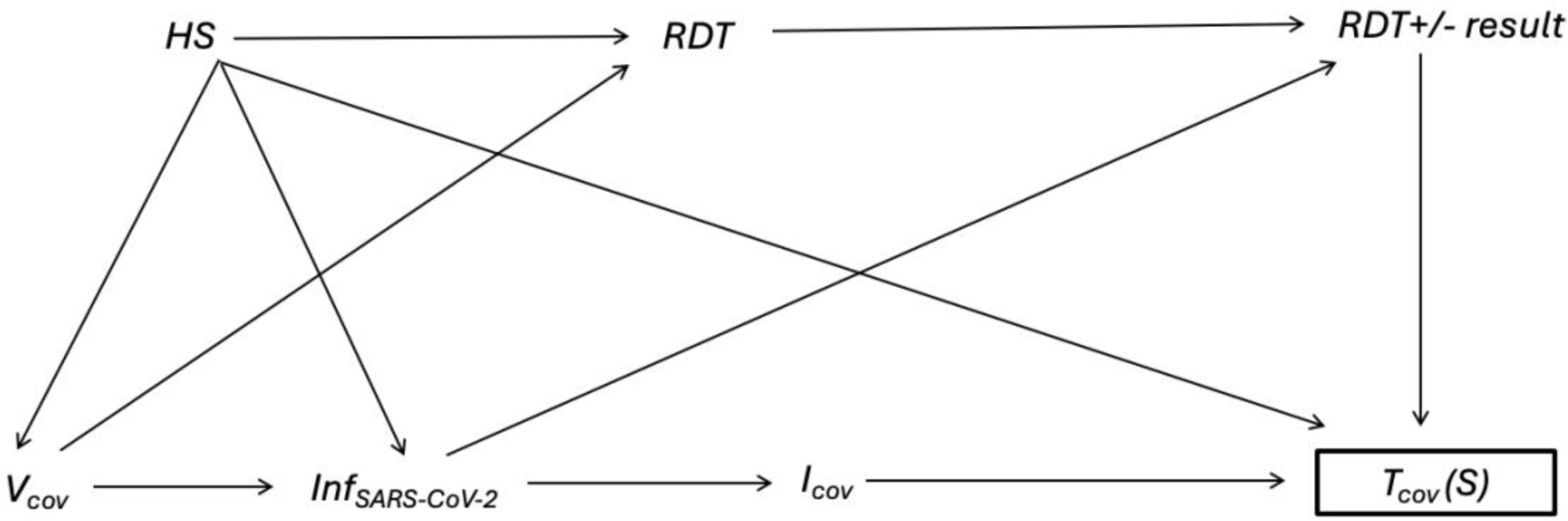
Directed acyclic graph (DAG) depicting causal relationships between at-home SARS-CoV-2 rapid diagnostic testing (*RDT*), home test result (*RDT result* [positive or negative]), COVID-19 vaccination status (*V_cov_*), SARS-CoV-2 infection (*Inf_SARS-CoV-2_*), COVID-19-like illness (*I_cov_*), healthcare seeking behavior (*HS*), and COVID-19 testing by a healthcare provider (*T_cov_*). The box around *T_cov_* (S) represents selection into the TND study and determination of COVID-19 case status. The DAG assumes all patients presenting with COVID-19-like illness symptoms are systematically tested for COVID-19 using molecular testing with high sensitivity and specificity. Other potential confounding factors including patient characteristics associated with *V_cov,_ HS, and I_cov_* are not depicted in this graph.

### Decision Tree Structure

We constructed a decision tree to simulate numbers of persons vaccinated and unvaccinated against COVID-19 experiencing acute respiratory illness (ARI) due to COVID-19 or non-COVID-19 illness who sought care and were subsequently included in the TND study as test-positive cases and test-negative control patients (Supplemental Figure 1). Individuals reporting ARI symptoms who were vaccinated are denoted by the probability *V_cov_,* or unvaccinated with probability 1-*V_cov_*. Probabilities of vaccinated and unvaccinated individuals taking at-home SARS-CoV-2 RDT for ARI symptoms are denoted by *RDT_v_* and *RDT_u_*, respectively. Probabilities of vaccinated and unvaccinated individuals seeking healthcare for ARI symptoms are denoted by *HS_v_* and *HS_u_,* respectively. *HS_v_RDT+*, *HS_v_RDT-*, *HS_u_RDT+*, and *HS_u_RDT-* denote probabilities of vaccinated and unvaccinated individuals with ARI seeking care following their at-home rapid diagnostic test result (positive or negative) (Table 1).

**Table 1:**
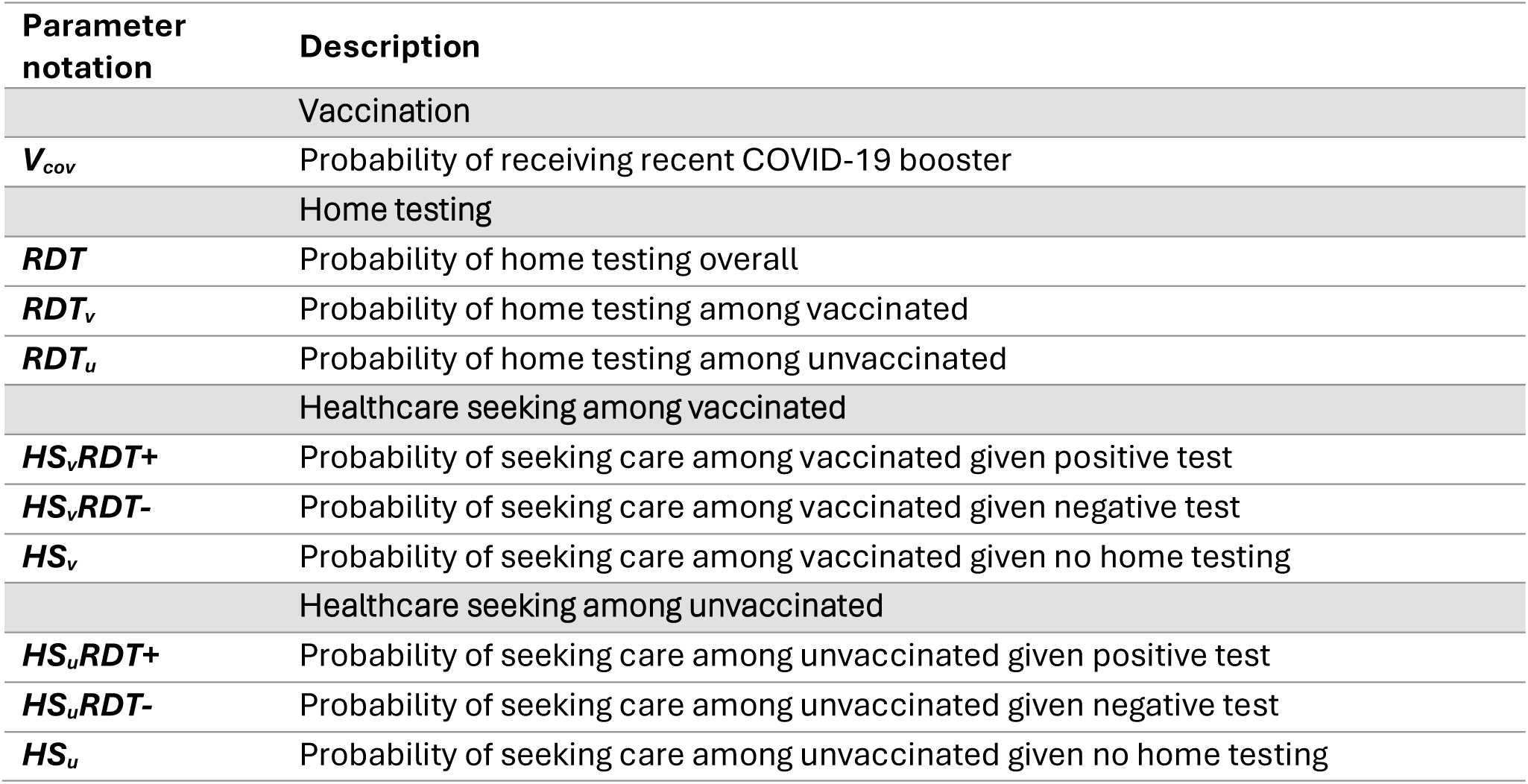
Parameters of factors that influence COVID-19 vaccine effectiveness estimation in symptomatic individuals sampled in test-negative design case-control studies.

As the effect of interest was the probability of seeking care for ARI symptoms based on at-home RDT test result rather than test accuracy among vaccinated and unvaccinated individuals, we assumed perfect concordance between the at-home RDT and provider-based testing that determine case status. All persons seeking care were assumed to be tested for SARS-CoV-2 infection and included in the TND study as SARS-CoV-2 test-positive cases or test-negative controls.

### Decision Tree Input Values

We varied the values of factors that could affect observed VE (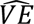), including COVID-19 vaccination uptake, probability of at-home RDT use that was stratified by vaccination status, and likelihood of seeking healthcare for ARI symptoms following home testing, also stratified by vaccination status and at-home RDT result. Each of these variables was defined by a probability distribution—a summary of the possible values of the variable—based on results from a national survey as described below. We used a beta distribution to represent each probability entered into the decision tree model. A beta distribution is typically used to represent probabilities because it is a flexible, continuous distribution bounded by values of zero and one^11^. Each beta distribution is summarized by two shape parameters: when representing the probability of a binary event, these parameters represent the number of positive responses observed and the number of negative responses observed (Supplemental Table 1). We obtained empirical values of these parameters directly from the CHASING COVID Cohort,^12^ a geographically and socio-demographically diverse longitudinal cohort study of approximately 6,740 adults residing in the U.S. and its territories. Cohort participants were recruited via social media or by referral and received electronic survey questionnaires approximately every three months beginning in March 2020. The surveys captured information on SARS-CoV-2 infections, COVID-19 testing behaviors, test results, vaccination status and healthcare seeking among other outcomes. Study methods and survey instruments are publicly available^12^. All CHASING COVID Cohort participants provided written informed consent before cohort enrollment. The IRB of the City University of New York gave ethical approval for the study procedures and protocols.

Probabilities were calculated using responses from participants who completed a survey between March 2022 – October 2023 and reported experiencing new ARI symptoms (cough, runny nose, sore throat, shortness of breath) since the previous survey. For participants with multiple episodes, only the first reported ARI was included. Vaccination status was defined based on reported receipt of the latest FDA-approved COVID-19 booster dose at the time of survey administration. For participants who completed the survey questionnaire administered between March 2022 and September 2022, vaccination was defined as those who received the mRNA monovalent COVID-19 booster vaccine (fourth COVID-19 dose). From September 2022 through September 2023, vaccination status was based as those who received the bivalent mRNA COVID-19 booster vaccine, and the updated 2023-2024 COVID-19 booster vaccine for participants completing the survey in October 2023. Unvaccinated participants were those who did not receive the most recent booster dose at or before prior survey’s fielding day. Probability of home testing was calculated as the proportion of participants reporting an ARI episode who took an at-home RDT because of acute symptoms. Participants who reported taking an at-home RDT during the time period for other reasons (e.g., required COVID-19 screening in the workplace) were classified as non-home testers. COVID-19 case status was based on a participant’s self-reported SARS-CoV-2 positive or negative viral diagnostic test for their ARI episode. Probability of seeking care for ARI was estimated from participants who reported seeing or calling a physician or health care professional for their ARI symptoms. Questions used to calculate each probability are shown in Supplemental Table 2.

In addition to the base model, we varied the decision tree parameters to reflect alternate scenarios by the following socio-demographic characteristics: race/ethnicity (non-Hispanic White, non-Hispanic Black, and Hispanic), education attainment (below, at or above high school attainment), and annual household income level (<$50,000, $50,000 – $100,000, and >$100,000).

### Simulations

Using Monte Carlo methods^11^, we simulated the total number of COVID-19 cases and controls, number of vaccinated cases and controls, and ratio of vaccinated to unvaccinated cases and controls based on the input probability distributions. The observed mean VE value (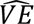, expressed as percent) was calculated as (1 – Odds Ratio [OR]) x100 comparing vaccination odds among test-positive cases versus test-negative controls. Credible intervals were defined by the 2.5^th^ and 97.5^th^ percentiles of the distribution of 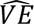. Percent bias was defined as the absolute difference between true *VE* and 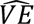. Simulations were run at six values of true *VE* (5, 10, 20, 40, 60, 80, 95) to assess magnitude of bias across a plausible range of COVID-19 VE. We varied values across the range of their distributions to examine the sensitivity of the results to changes in parameter values. Tornado plots were used to examine the sensitivity 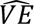 by varying values of each parameter by +/- one standard deviation (SD) from the mean. We show in descending rank order the most to least influential parameters that effect estimated 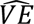. At true *VE* set at high (60%) and low (20%) values, the input value of the most influential parameter was varied by +/- 10% from the mean to examine the influence on 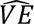. We further assessed magnitude of bias across socio-demographic characteristics with true *VE* set at 20% and 60%. Finally, we performed two sensitivity analyses at a true VE of 20%: 1) simulations were conducted with an at-home RDT sensitivity of 64% and 100% specificity, based on a review of RDT sensitivity^13^; and 2) adjusting proportions seeking care after a positive test if the at-home RDT was negative. For the second sensitivity analysis, we used responses from one survey round among symptomatic participants who reported taking an at-home RDT, seeking care and testing positive with a provider (Supplementary Table 2). Of these, 24% reported testing positive after seeking care. VE simulations were conducted using @RISKv8.6 (Palisade Corp, Ithaca, NY).

## Results

Input probabilities for the base decision tree model were obtained by summarizing responses from 1,918 participants with ARI symptoms between March 2022 – October 2023 (Table 2). Overall, 70% of participants in the study sample during that period reported receiving recent COVID-19 vaccination. A total of 33% of participants with ARI symptoms reported use of an at-home RDT for their illness: 37% of vaccinated and 22% of unvaccinated participants. Healthcare seeking for ARI was associated with COVID-19 vaccination and at-home RDT use. Among vaccinated participants, 47% of those reporting a positive at-home test, 37% of those reporting a negative test, and 23% of those who did not take an at-home RDT sought care for their ARI symptoms. Among unvaccinated participants, 44% of those reporting a positive test, 24% of those reporting negative test, and 23% of those who did not use an at-home RDT sought care for their ARI symptoms (Table 2).

**Table 2:**
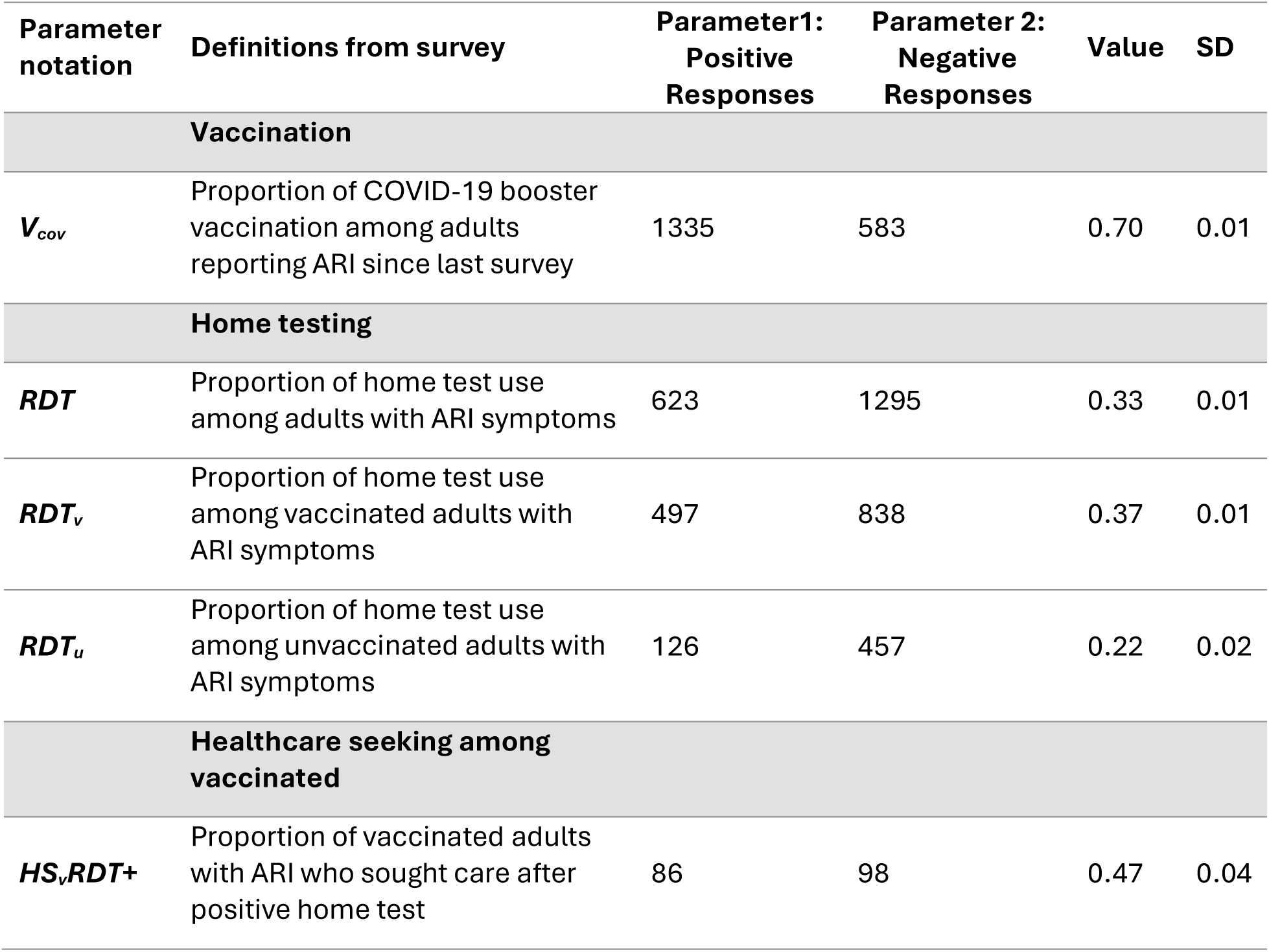
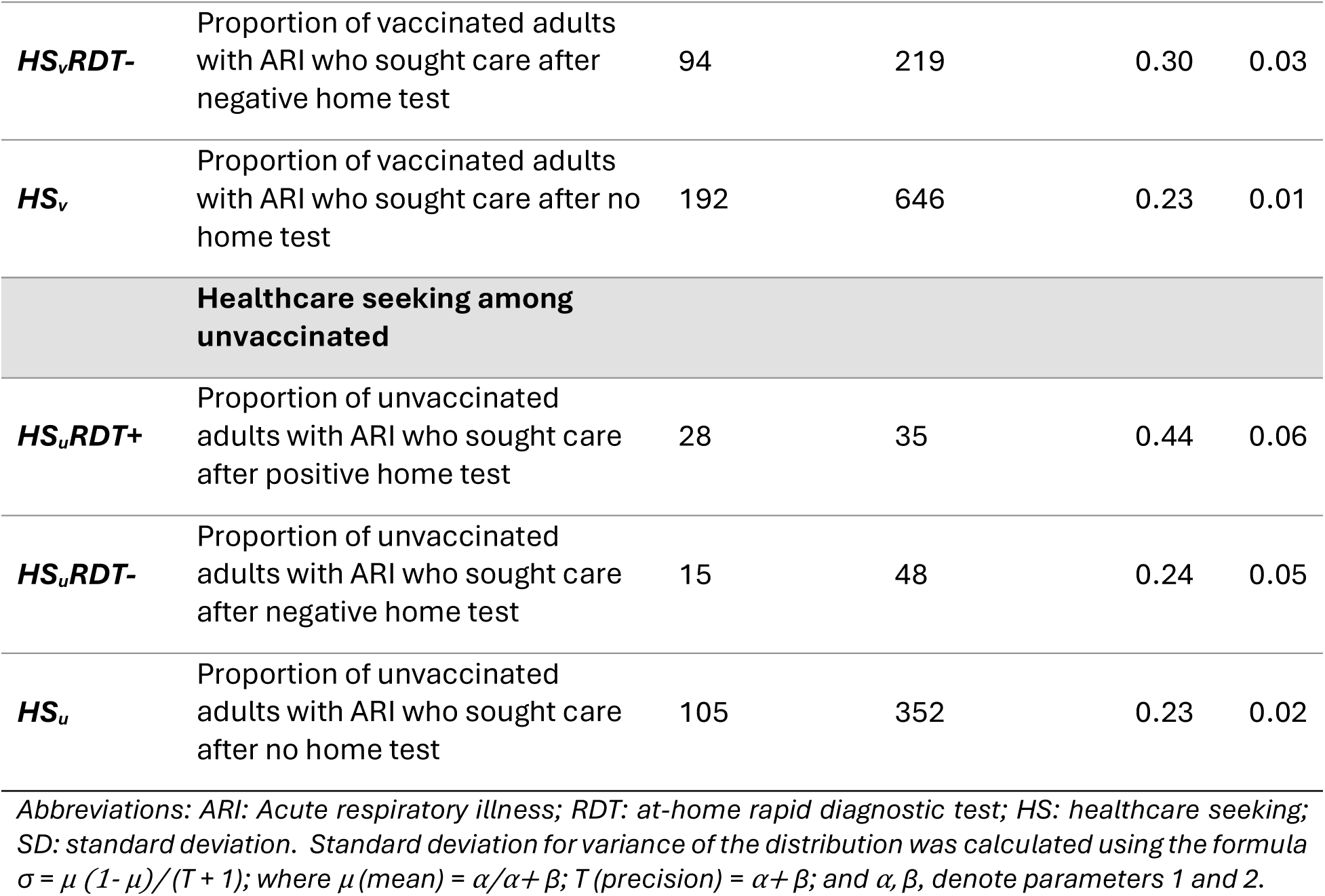
Decision tree probability values based on survey responses among participants reporting acute respiratory illness symptoms during March 2022 - October 2023 (N = 1,918)

In the base model with 95% credible intervals obtained from Monte Carlo simulations, 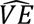 underestimated true *VE* in all model iterations with increasing bias at lower levels of true *VE*. The decision tree model simulations with input values from the survey data indicated a percent bias of 5-percentage point downward difference between 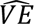 and true *VE* value at values from 5% to 95%. This downward bias was minimal at true *VE* above 40% and approached 5-percentage point difference at true *VE* below 40%. At true *VE* of 20%, 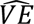 was 16.3%. At true *VE* of 60%, 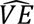 was 58.1% (Figure 2).

**Figure 2:**
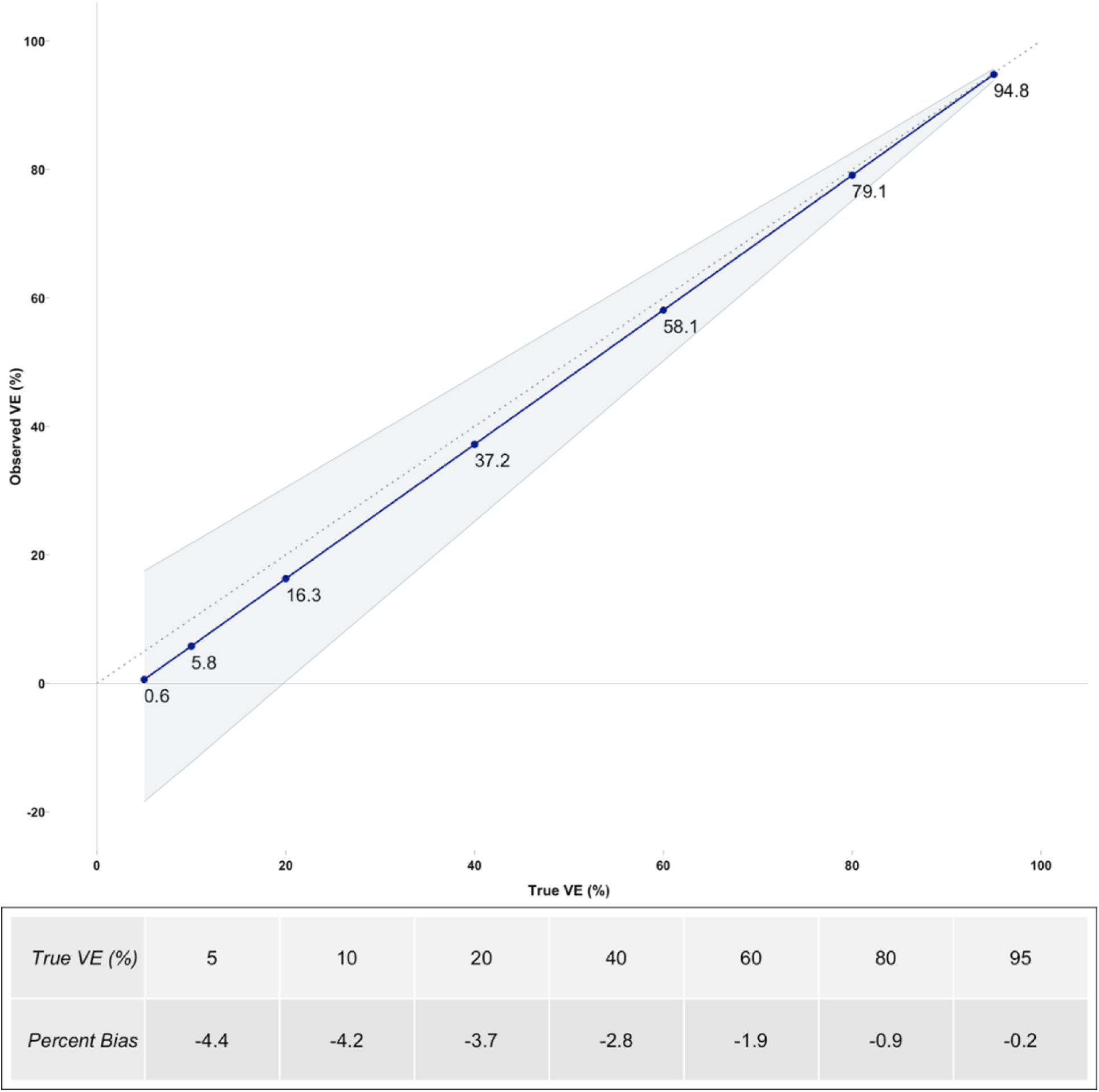
G*r*aph *showing simulated observed vaccine effectiveness (*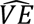*) (blue line) against ranging true VE values (5, 10, 20, 40, 60, 80, 95) in the base model. The gray shaded area represents the 95% credible intervals. The dotted gray line is the line of identity (*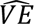 *= VE*)*. Percent bias is the absolute difference between* 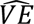 *and true VE*.

The magnitude but not the rank order of influence of a parameter on estimated VE varied by true *VE* values. Probability of healthcare seeking among unvaccinated adults reporting negative at-home RDT, *HS_u_RDT-*, and the probability of at-home RDT use among vaccinated adults, *RDT_v_*, were the most and least influential parameters on VE estimates, respectively (Figure 3). At a true *VE* of 20%, one SD higher in the probability of healthcare seeking among unvaccinated adults who tested positive resulted in a 4-percentage point increase in 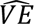 (Figure 3a) and 2-percentage point increase in 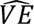 when true *VE* was 60% (Figure 3b).

**Figure 3:**
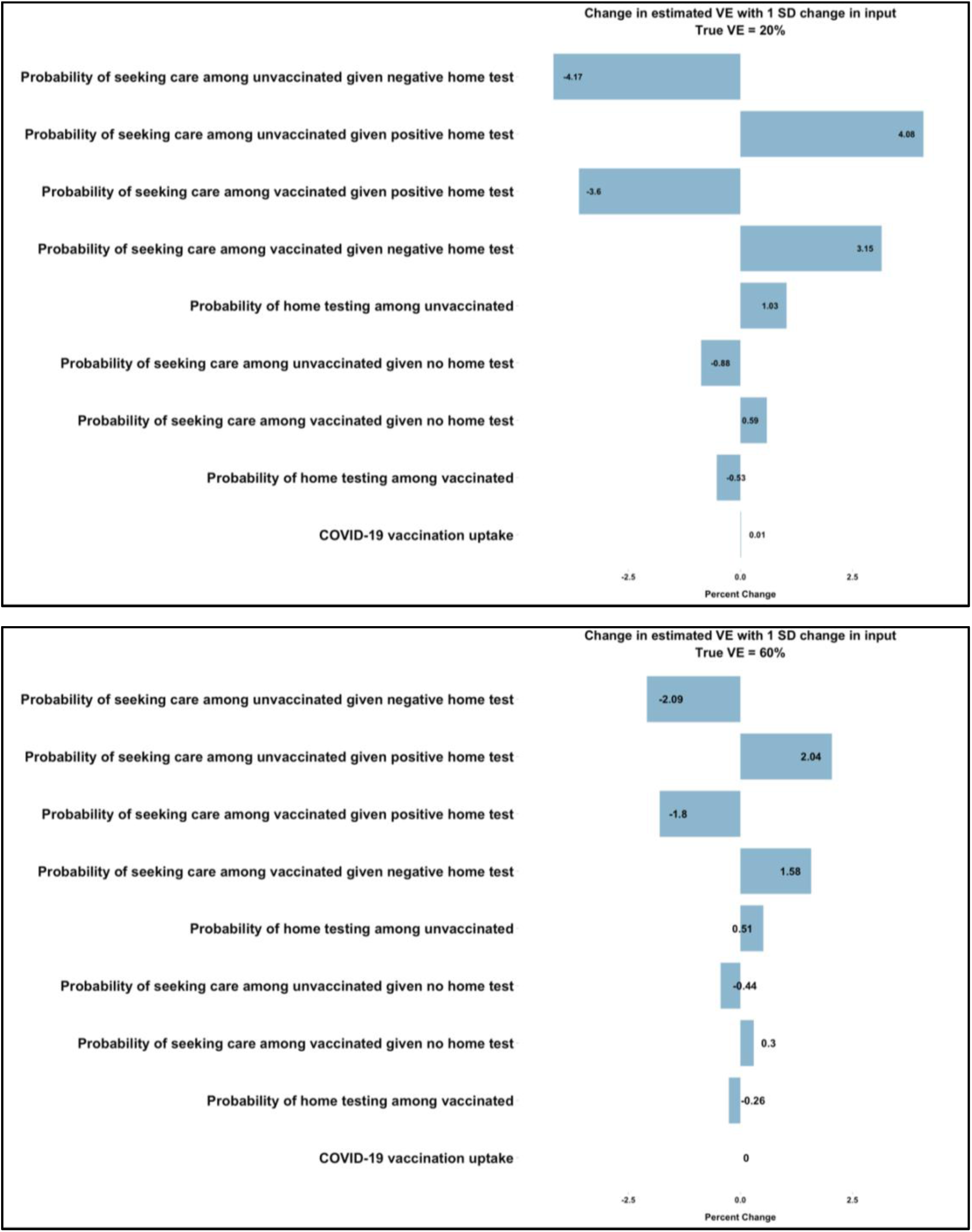
Tornado plot showing the effects of varying parameters on 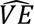. Plots show in rank order the most to least (top-down y-axis) influential parameters that affect estimated 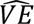. Figure 3a (top plot) shows the effect of one standard deviation increase in each parameter on change in 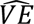 when true VE is 20%. Figure 3b (bottom plot) shows effects of one standard deviation increase in each parameter on change in 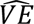 when true VE is 60%. These values can be interpreted as the percentage point change for each one standard deviation increase in the respective model input.

To examine the effect of varying the most influential variable on downward bias in 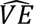, probability of seeking care among unvaccinated adults with ARI following negative at-home RDT, we raised the input probability for *HS_u_RDT-* by 10% to 0.34. When true *VE* was below 10%, estimated 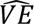 was negative. At true *VE* of 20% or 60%, 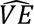 was 8.5% and 77.1%, respectively. Alternatively, an input probability of 0.14 for *HS_u_RDT-,* resulted in 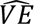 of 24.1% at true *VE* of 20%, and 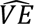 of 62.1% at true *VE* of 60% (Supplemental Table 3).

In sensitivity analyses with true *VE* of 20%, a lower at-home RDT sensitivity resulted in a 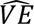 of 17.6%. A higher proportion of persons seeking care following a negative at-home RDT resulted in a 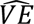 of 17.5% when true VE was 20%.

Finally, we examined the effect on 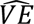 by replacing input probabilities from the entire survey sample with probabilities based on socio-demographic characteristics of survey participants. Overall vaccination uptake *V_cov_* and probability of at-home *RDT* use varied by socio-demographic characteristics (Table 3). However, at-home RDT use among vaccinated participants, *RDT_v_* was consistently higher than among unvaccinated participants, *RDT_u_*. Differences in the magnitude and direction of bias in 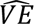 illustrated the influence of multiple factors. Among participants reporting annual household income >$100,000, a high *V_cov_*(0.87) and at-home *RDT* use (0.42), combined with a lower probability of healthcare seeking following a negative *RDT* among vaccinated, *HS_v_RDT- (*0.24), compared to unvaccinated, *HS_u_RDT- (*0.40), resulted in 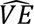 of -22% at true *VE* of 20% and 39% at true *VE* of 60%. In contrast, among non-Hispanic Black participants and those reporting annual household income between $50,000-$100,000, higher probability of healthcare seeking following a positive test among unvaccinated, *HS_u_RDT+*, versus vaccinated participants, *HS_v_RDT+*, resulted in a substantially overestimated 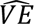.

**Table 3:**
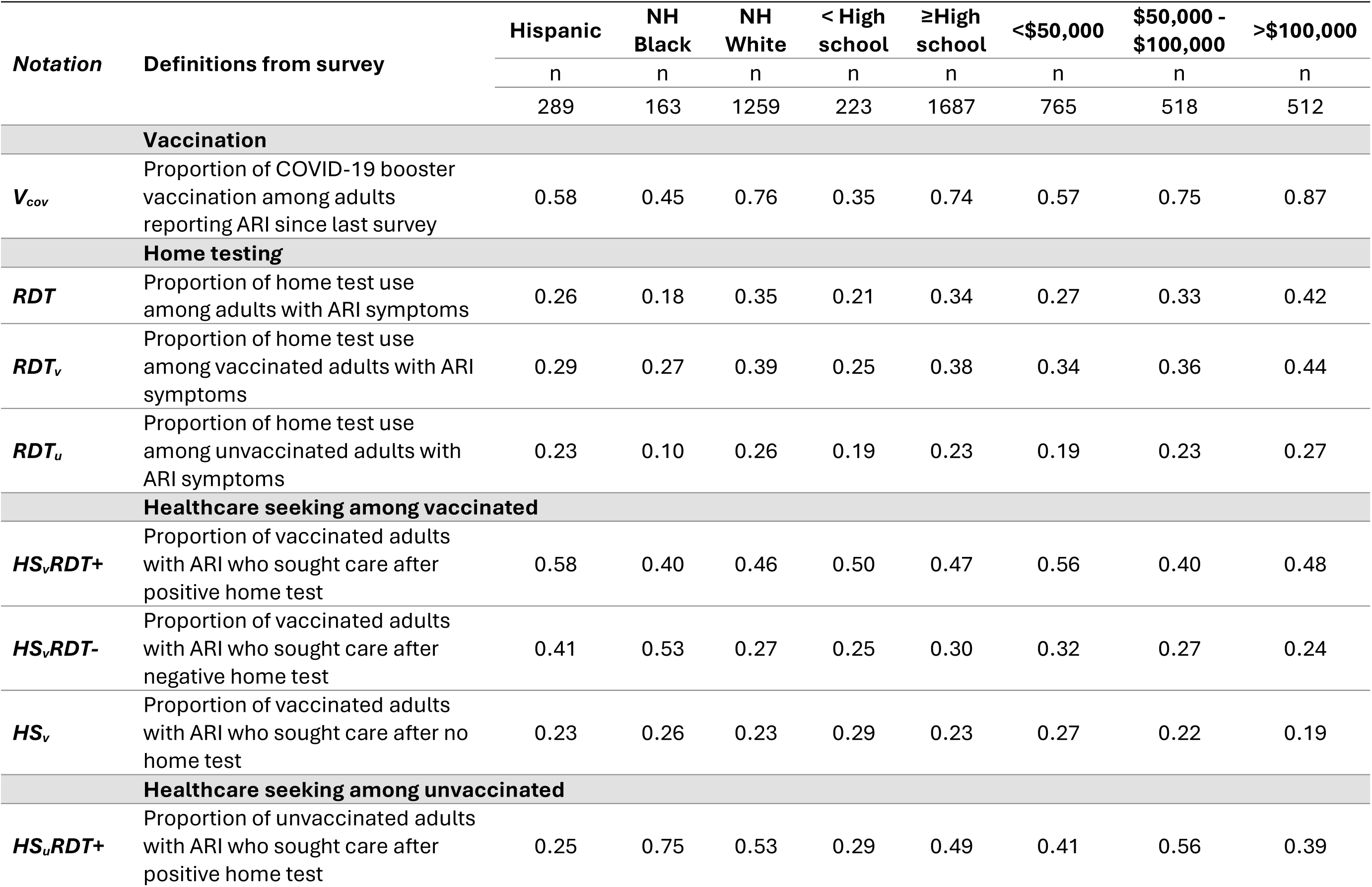
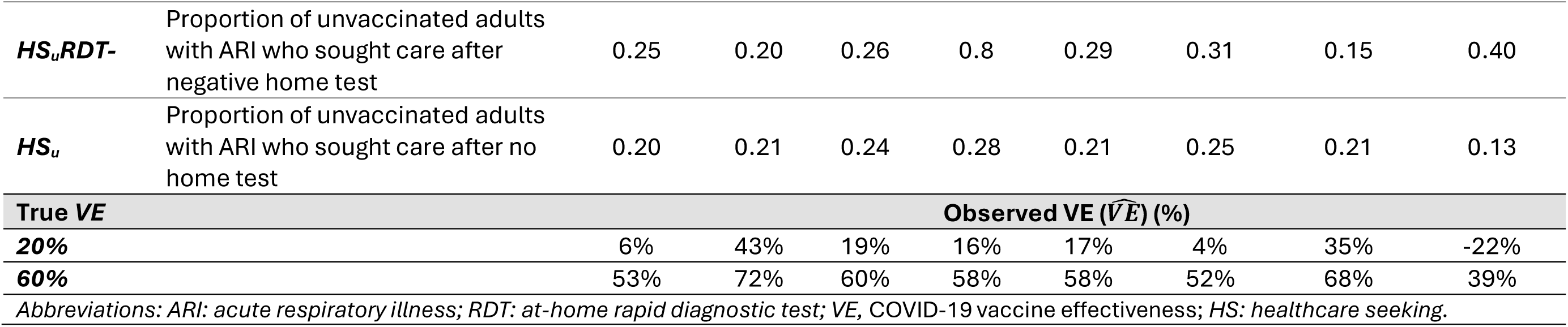
Decision tree input values for factors associated with COVID-19 vaccine effectiveness estimation across select socio-demographic characteristics, March 2022 – October 2023, (N=1,918)

## Discussion

Our study presents a theoretical model to assess potential bias in estimates of COVID-19 vaccine effectiveness using TND case-control studies when at-home RDT use influences healthcare seeking for COVID-19-like illness. Using probabilities for key input parameters based on a diverse group of survey respondents in a longitudinal cohort, observed COVID-19 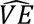 underestimated true *VE* by less than 5 percentage points across a range of true *VE* values. Our observed bias was within confidence bounds of many COVID-19 vaccine effectiveness estimates^14^. However, by substituting probabilities observed in specific socio-demographic groups of participants, we found potentially large biases in either direction. Differences in healthcare seeking among vaccinated versus unvaccinated persons following home testing influenced the magnitude and direction of bias. In contrast, there is less potential for bias from variation in healthcare seeking behavior among persons who did not home test, based on probabilities derived from survey data.

Importantly, based on probabilities of COVID-19 vaccination, home testing and healthcare seeking observed among survey respondents with annual household incomes >$100,000, apparent 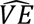 estimates were negative at low values of vaccine effectiveness. While we observed minimal bias when vaccination was >80% effective, we observed downward bias resulting in null or negative findings at true VE <40%. Findings of null or negative VE findings may contribute to lack of public confidence that vaccine provides benefit, potentially depressing vaccine demand and uptake^15,16^. Among respondents with household incomes >$100,000 and vaccination uptake >70%, underestimation of 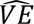 was associated with higher percentages reporting at-home RDT for illness and seeking care following positive RDT results. Among non-Hispanic Black participants, overestimation of 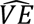 was associated with higher probability of healthcare seeking among unvaccinated cases. Knowledge of at-home RDT use among populations seeking care is important when interpreting apparent differences in vaccine effectiveness by socio-demographic characteristics of patients.

In simulations, we observed a potential for selection bias in TND studies when home testing influenced symptom-driven healthcare seeking by vaccination status among cases and controls. In our base model, the probability of healthcare seeking following a test-positive result was higher among vaccinated than unvaccinated persons, and lower among vaccinated test-negative persons, favoring the selection of vaccinated cases and resulting in underestimated vaccine effectiveness. When vaccination is highly protective, vaccinated persons would be less likely to have COVID-19 and test positive by RDT if they took an at-home test, resulting in less bias even when healthcare seeking is influenced by at-home RDT use. In simulations, lower RDT sensitivity also reduced VE bias because participants were less likely to seek care after a negative test.

The observed higher probability of healthcare seeking among vaccinated persons using at-home SARS-CoV-2 RDT may be explained by general health protective behaviors among those who received the COVID-19 vaccine^5^. At-home SARS-CoV-2 RDT use has been shown to be associated with vaccination status^17^ and higher SES^10^. In addition, healthcare access and use of antivirals such as nirmatrelvir/ritonavir (Paxlovid^TM^) have been shown to have disparities across race and ethnicity^18^.

Our assessment of bias based on differential probabilities across socio-demographic characteristics highlighted the impact of differences in at-home RDT use and healthcare seeking on VE estimation from TND studies conducted in different populations. Care settings are associated with patient socioeconomic characteristics^19,20^. For example, insured patients who are more likely to be vaccinated and test at home may be overrepresented in outpatient settings, whereas uninsured and under-insured patients may make up a larger proportion of hospitalized patients^21^ highlighting the potential implications on TND studies and VE estimation.

Testing using over the counter at-home RDTs for COVID-19, influenza and respiratory syncytial virus is likely to become more widely available. Monitoring at-home RDT uptake is needed to evaluate potential influence on vaccine effectiveness studies for these vaccine preventable respiratory diseases^22^. Previous studies have demonstrated the potential underestimation of COVID-19 incidence when results of at-home SARS-CoV-2 RDT were not included in case counts^23^. In 2021, almost half of CHASING COVID cohort participants who reported use of at-home RDT for COVID-19-like symptoms had used at-home tests exclusively and did not receive testing from a provider. Significant underestimation of COVID-19 prevalence was also observed in comparison to estimates from population-representative adults in NYC and in the U.S^24,25^. High uptake of at-home RDT can not only underestimate infection prevalence, but can lead to potential bias in VE studies and impact communication of the benefits of the vaccination ^22^.

Our study had several limitations. First, we used probabilities for input parameters from a non-representative cohort of survey respondents. Second, survey data were collected over a 20-month period when home tests were widely available through federal and local governments^26,27^. We did not account for temporal trends in healthcare seeking and use of at-home SARS-CoV-2 RDT. Third, we assumed the at-home SARS-CoV-2 RDT result was the same as provider-test for TND case classification. Among a subset of participants asked specifically about the result for at-home RDT, magnitude and direction of bias were similar to the base model assuming that a proportion of non-COVID-19 patients had a false positive RDT that influenced their decision to seek healthcare. Fourth, in the main analysis we assumed there were no differences in at-home RDT sensitivity and specificity by vaccination status or prior infection. The use of less sensitive at-home RDTs tended to reduce the magnitude but not change the direction of bias. Finally, COVID-19 vaccination, testing and healthcare behaviors were based on self-report which may not reflect true behaviors.

This study highlights the potential impact of at-home SARS-CoV-2 RDT use on COVID-19 vaccine effectiveness estimation. Population-based surveys are needed to monitor uptake of home testing^28,29^; other sources include electronic medical records or manufacturer supply chain information^30^. Information about healthcare seeking and COVID-19 home testing is needed to evaluate potential for biased estimates from observational studies using TND.

## Data Availability

All data produced in the present study are available upon reasonable request to the authors.

---

**Supplemental Figure 1:**
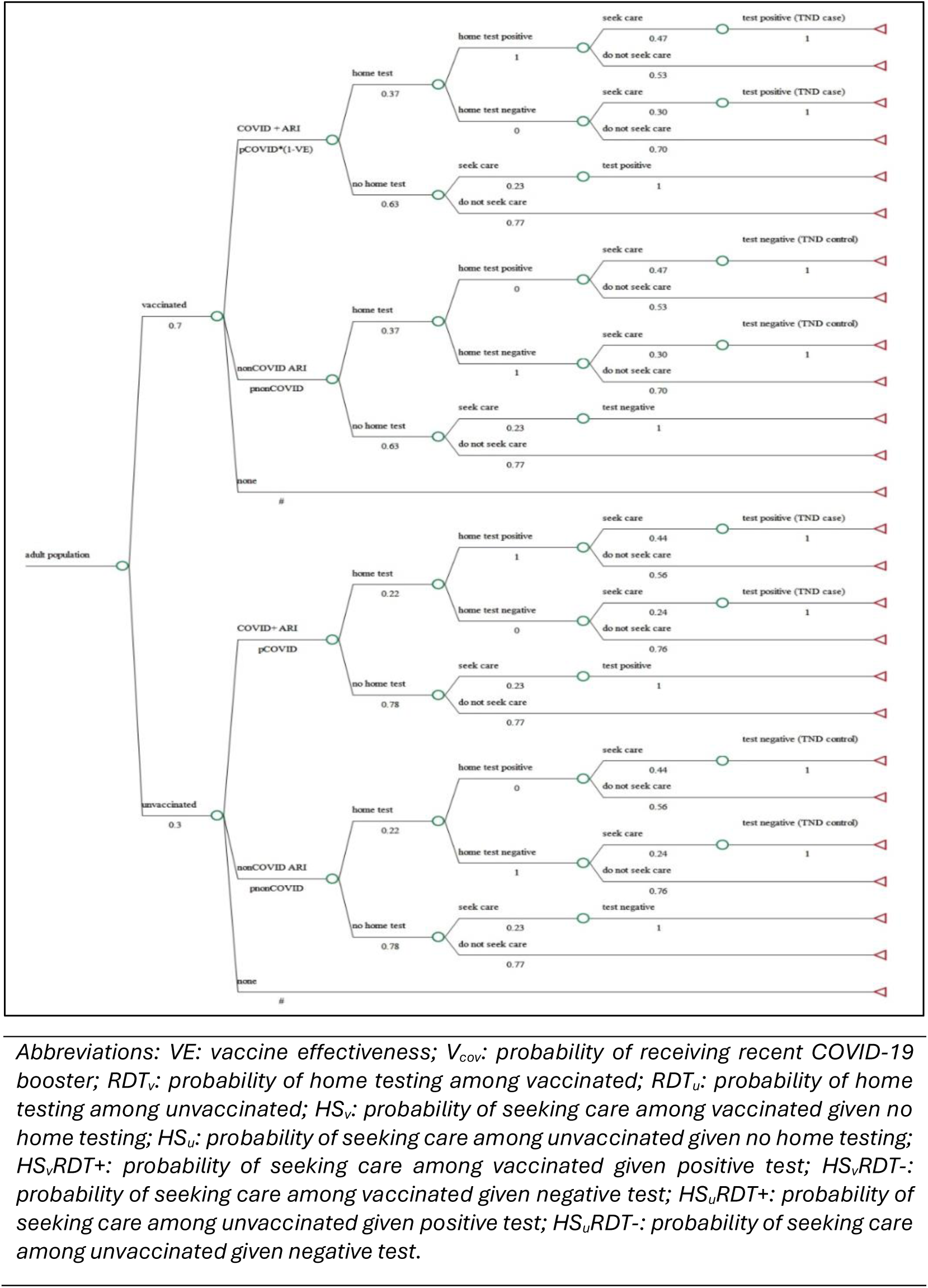
Decision tree depicting factors associated with COVID-19 vaccine effectiveness estimation. Chance nodes are shown by circles, which denote outcomes that may occur by chance at each point in the tree with likelihood of occurrence given by the probability shown under each branch (*V_cov_, RDT_v_*, *RDT_u_, HS_v_*, *HS_u_*, *HS_v_RDT+*, *HS_v_RDT-*, *HS_u_RDT+*, and *HS_u_RDT-*). Terminal nodes (triangles) represent the observed study outcomes of cases and controls based on healthcare provider SARS-CoV-2 test result (positive or negative).

**Supplemental Table 1:**
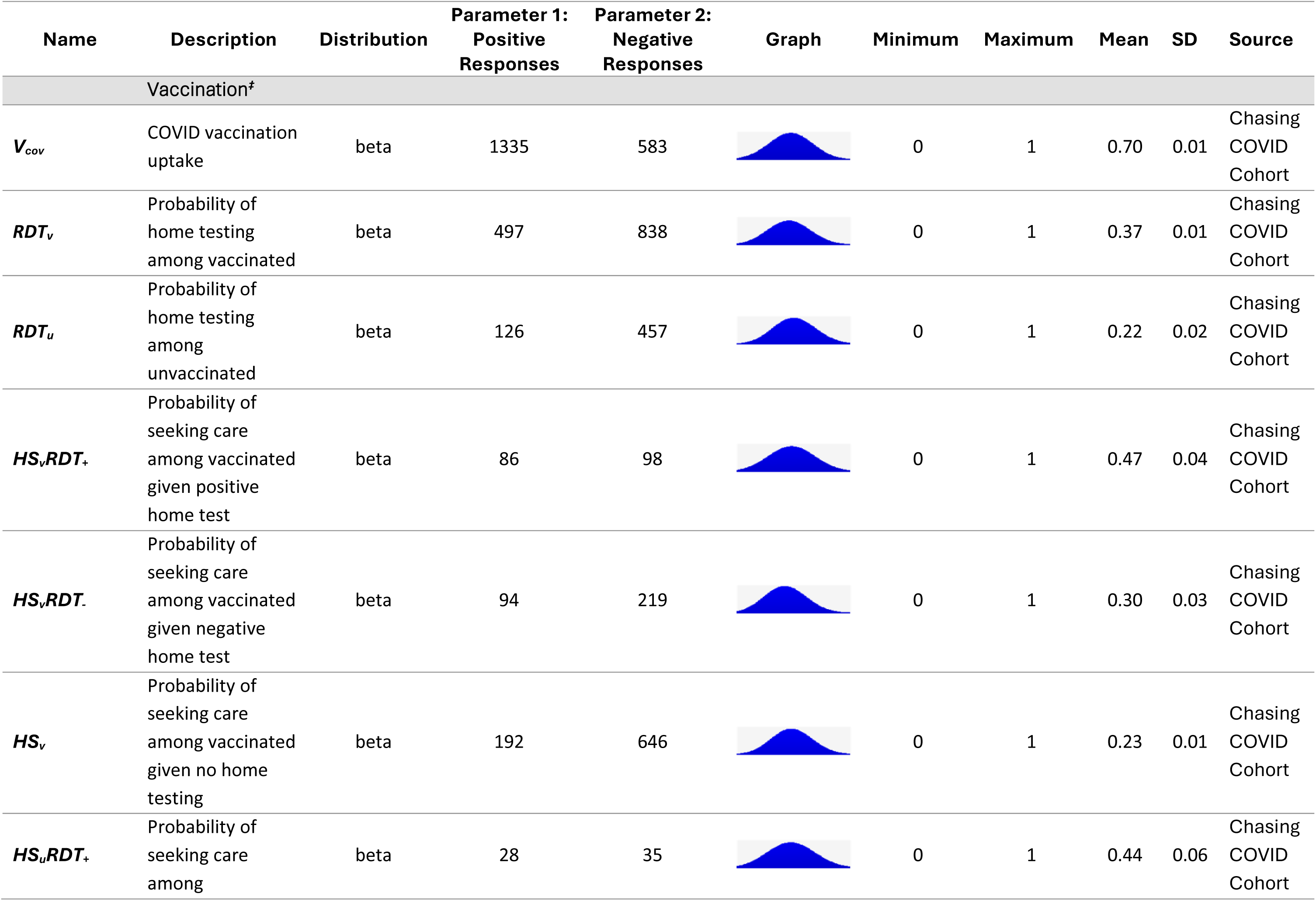
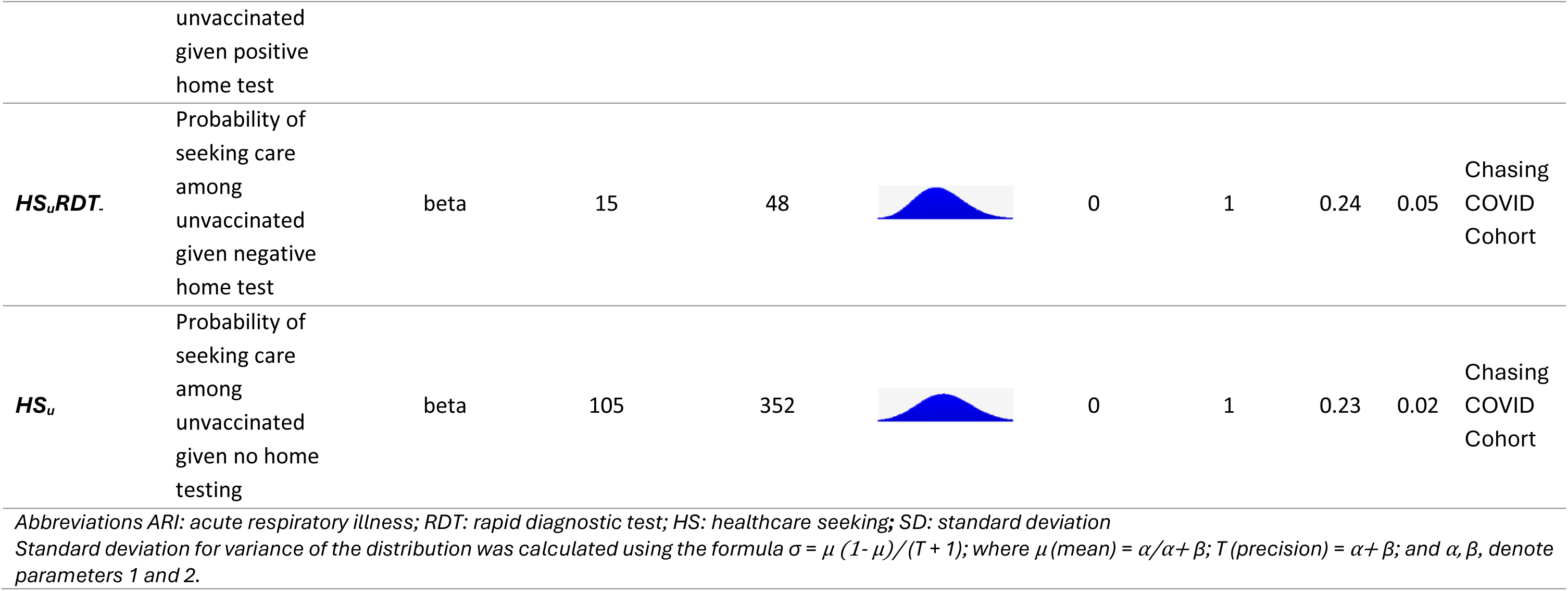
Beta distribution shape parameters for decision tree factors affecting COVID-19 vaccine effectiveness estimation.

**Supplemental Table 2:**
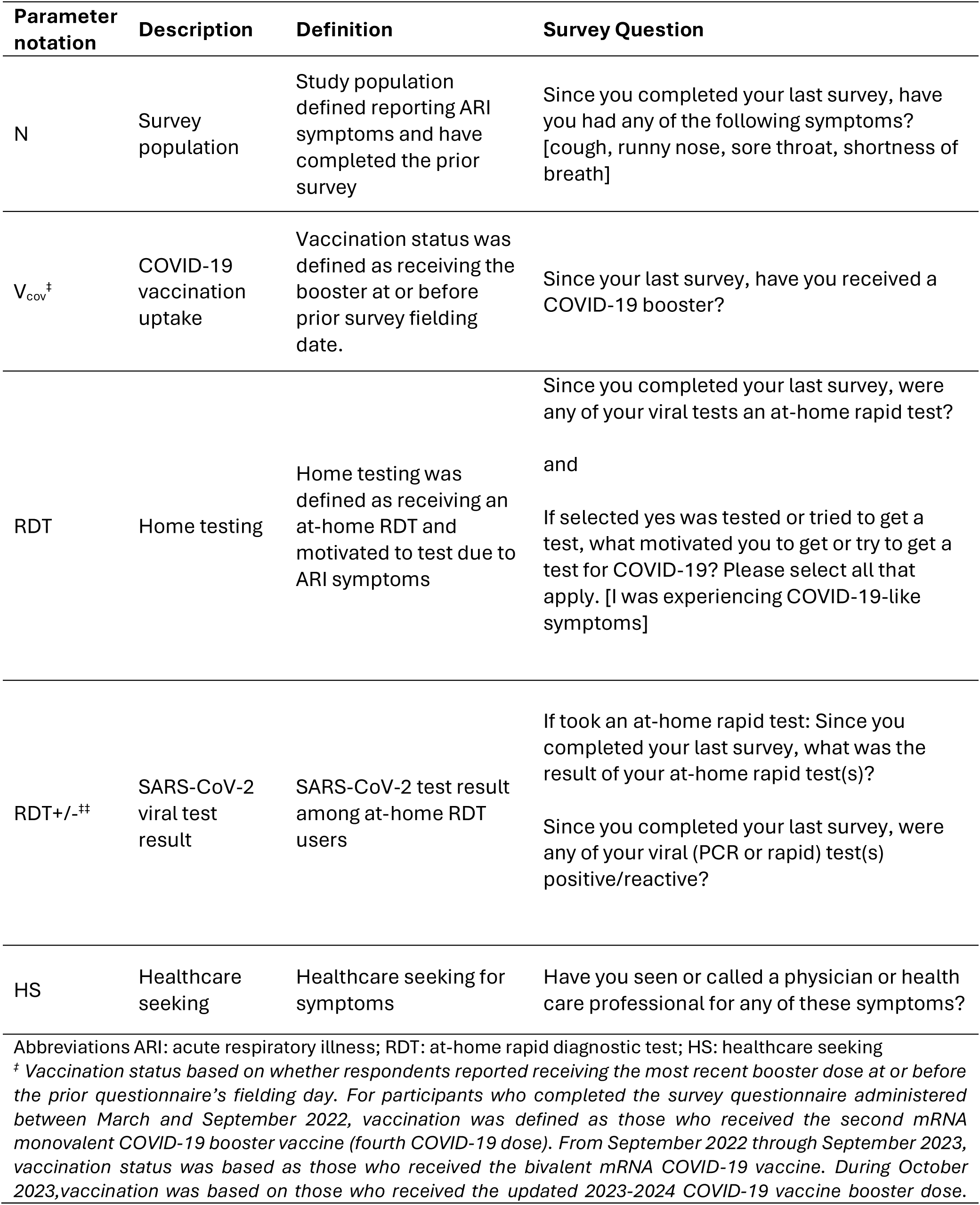
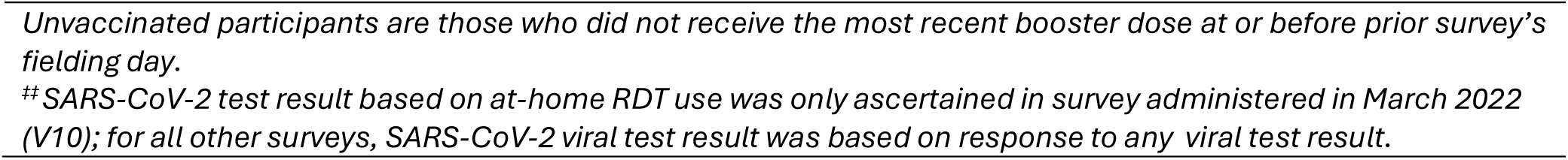
Definition of simulation parameters from CHASING COVID Cohort study surveys.

**Supplemental Table 3:** Bias between true *VE* and 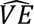 based on 10%-point increase or decrease of probability of healthcare seeking among unvaccinated test-negative controls in the base model.

| True VE (%) | Probability of healthcare seeking among unvaccinated controls ($HS_uRDT$-) | | | |
| --- | --- | --- | --- | --- |
| | + 10%-point $HS_uRDT$- | Percent bias<sup>‡</sup> | - 10%-point $HS_uRDT$- | Percent bias<sup>‡</sup> |
| 5 | -8.7 | -13.7 | 9.9 | 4.9 |
| 10 | -3.0 | -13.0 | 14.6 | 4.6 |
| 20 | 8.5 | -11.5 | 24.1 | 4.1 |
| 40 | 31.3 | -8.7 | 43.1 | 3.1 |
| 60 | 54.2 | -5.8 | 62.1 | 2.1 |
| 80 | 77.1 | -2.9 | 81.0 | 1.0 |
| 95 | 94.28 | -0.7 | 95.26 | 0.3 |

<sup>‡</sup>Percent bias is the absolute difference between observed VE ( $\widehat{VE}$ ) and true VE.
Abbreviations ARI: acute respiratory illness; RDT: rapid diagnostic test; HS: healthcare seeking; VE: vaccine COVID-19 effectiveness

## References

1. Pouwels KB, Pritchard E, Matthews PC, et al. Effect of Delta variant on viral burden and vaccine effectiveness against new SARS-CoV-2 infections in the UK. Nat Med. 2021;27(12):2127–2135.

2. Payne AB, Ciesla AA, Rowley EAK, et al. Impact of accounting for correlation between COVID-19 and influenza vaccination in a COVID-19 vaccine effectiveness evaluation using a test-negative design. Vaccine. Published online November 23, 2023. doi:10.1016/j.vaccine.2023.11.025

3. Dean NE, Hogan JW, Schnitzer ME. Covid-19 vaccine effectiveness and the test-negative design. N Engl J Med. 2021;385(15):1431–1433.

4. Jackson ML, Nelson JC. The test-negative design for estimating influenza vaccine effectiveness. Vaccine. 2013;31(17):2165–2168.

5. Sullivan SG, Tchetgen Tchetgen EJ, Cowling BJ. Theoretical basis of the test-negative study design for assessment of influenza vaccine effectiveness. Am J Epidemiol. 2016;184(5):345–353.

6. Ciocănea-Teodorescu I, Nason M, Sjölander A, Gabriel EE. Adjustment for disease severity in the test-negative study design. Am J Epidemiol. 2021;190(9):1882–1889.

7. Foppa IM, Haber M, Ferdinands JM, Shay DK. The case test-negative design for studies of the effectiveness of influenza vaccine. Vaccine. 2013;31(30):3104–3109.

8. Goggolidou P, Hodges-Mameletzis I, Purewal S, Karakoula A, Warr T. Self-testing as an invaluable tool in fighting the COVID-19 pandemic. J Prim Care Community Health. 2021;12:21501327211047784.

9. Standing Committee for CDC Center for Preparedness and Response, Health and Medicine Division, National Academies of Sciences, Engineering, and Medicine. Rapid Expert Consultation on Self-Tests for Infectious Diseases. National Academies Press; 2022.

10. Rader B, Gertz A, Iuliano AD, et al. Use of At-Home COVID-19 Tests - United States, August 23, 2021-March 12, 2022. MMWR Morb Mortal Wkly Rep. 2022;71(13):489–494.

11. Vose D. A Guide to Monte Carlo Simulation Modeling. Wiley; 1996.

12. CHASING COVID cohort study. CUNY ISPH. Accessed March 17, 2024. https://cunyisph.org/chasing-covid/

13. Prince-Guerra JL, Almendares O, Nolen LD, et al. Evaluation of Abbott BinaxNOW rapid antigen test for SARS-CoV-2 infection at two community-based testing sites - Pima County, Arizona, November 3-17, 2020. MMWR Morb Mortal Wkly Rep. 2021;70(3):100–105.

14. Wiegand R, Fireman B, Najdowski M, Tenforde M, Link-Gelles R, Ferdinands J. Bias and negative values of COVID-19 vaccine effectiveness estimates from a test-negative design without controlling for prior SARS-CoV-2 infection. Research Square. Published online August 5, 2024. https://www.researchsquare.com/article/rs-4802667/v1

15. Weinstein N, Schwarz K, Chan I, et al. COVID-19 vaccine hesitancy among US adults: Safety and effectiveness perceptions and messaging to increase vaccine confidence and intent to vaccinate. Public Health Rep. 2024;139(1):102–111.

16. Nguyen KH, Chung EL, McChesney C, Vasudevan L, Allen JD, Bednarczyk RA. Changes in general and COVID-19 vaccine hesitancy among U.S. adults from 2021 to 2022. Ann Med. 2024;56(1):2357230.

17. Dorabawila V, Barnes V, Ramesh N, et al. Comparison of COVID-19 home-testers vs. laboratory-testers in New York State (excluding New York City), November 2021 to April 2022. Front Public Health. 2023;11:1058644.

18. Boehmer TK, Koumans EH, Skillen EL, et al. Racial and ethnic disparities in outpatient treatment of COVID-19 - United States, January-July 2022. MMWR Morb Mortal Wkly Rep. 2022;71(43):1359–1365.

19. Sparling A, Walls M, Mayfield CA, et al. Racial/ethnic disparities in health care setting choice for adults seeking severe acute respiratory syndrome Coronavirus 2 testing. Med Care. 2022;60(1):3–12.

20. Priem JS, Krinner LM, Constantine ST, McCurdy L. Diversification of COVID-19 testing resources to decrease racial/ethnic disparities: Comparative use of adaptive approaches to community testing across an integrated healthcare system. Dialogues Health. 2022;1(100017):100017.

21. Zhou RA, Baicker K, Taubman S, Finkelstein AN. The uninsured do not use the emergency department more-they use other care less. Health Aff (Millwood). 2017;36(12):2115–2122.

22. Bodner K, Irvine MA, Kwong JC, Mishra S. Observed negative vaccine effectiveness could be the canary in the coal mine for biases in observational COVID-19 studies. Int J Infect Dis. 2023;131:111–114.

23. Qasmieh SA, Robertson MM, Rane MS, et al. The importance of incorporating at-home testing into SARS-CoV-2 point prevalence estimates: Findings from a US national cohort, February 2022. JMIR Public Health Surveill. 2022;8(12):e38196.

24. Qasmieh SA, Robertson MM, Teasdale CA, et al. The prevalence of SARS-CoV-2 infection and other public health outcomes during the BA.2/BA.2.12.1 surge, New York City, April-May 2022. Commun Med (Lond). 2023;3(1):92.

25. Qasmieh SA, Robertson MM, Teasdale CA, et al. The prevalence of SARS-CoV-2 infection and long COVID in U.S. adults during the BA.4/BA.5 surge, June-July 2022. Prev Med. 2023;169(107461):107461.

26. Assistant Secretary for Public Affairs (ASPA). Biden-Harris Administration requires insurance companies and group health plans to cover the cost of at-home COVID-19 tests, increasing access to free tests. US Department of Health and Human Services. January 10, 2022. Accessed April 2, 2023. https://www.hhs.gov/about/news/2022/01/10/biden-harris-administration-requires-insurance-companies-group-health-plans-to-cover-cost-at-home-covid-19-tests-increasing-access-free-tests.html

27. The White House. Fact sheet: The Biden Administration to begin distributing at-home, rapid COVID-19 tests to Americans for Free. The White House. January 14, 2022. Accessed April 2, 2023. https://www.whitehouse.gov/briefing-room/statements-releases/2022/01/14/fact-sheet-the-biden-administration-to-begin-distributing-at-home-rapid-covid-19-tests-to-americans-for-free/

28. Qasmieh, S., Robertson, M. & Nash, D. “Boosting” surveillance for a more impactful public health response during protracted and evolving infection disease threats: insights from the COVID-19 pandemic. Health Security (in press). Health Secu.

29. Luisi N, Sullivan PS, Sanchez T, et al. Use of Covidtests.gov at-home test kits among adults in a national household probability sample - United States, 2022. MMWR Morb Mortal Wkly Rep. 2023;72(16):445–449.

30. Ritchey MD, Rosenblum HG, Del Guercio K, et al. COVID-19 Self-Test Data: Challenges and Opportunities — United States, October 31, 2021–June 11, 2022. MMWR Morbidity and Mortality Weekly Report. 2022;71(32):1005–1010. doi:10.15585/mmwr.mm7132a1

